# Barriers to care after Hepatitis C diagnosis: an implementation study of assisted self-testing among people who use drugs in Johannesburg

**DOI:** 10.64898/2026.02.05.26345695

**Authors:** Vanessa Tiyamike Msolomba, Yasmin Dunkley, Siphamandla Gumede, Mohammed Majam, Albert Manyuchi, Nomfundo Nhlapo, Karin Hatzold, Francois Venter

**Affiliations:** Faculty of Health Science, Clinical Medicine, Ezintsha, University of Witwatersrand, South Africa; Department of Clinical Research, Faculty of Infectious and Tropical Diseases. London School of Hygiene and Tropical Medicine, Botswana; Global HIV, TB and Viral Hepatitis Programs, Population Services International, South Africa; Anova Health institute, South Africa

**Keywords:** Hepatitis C, self-testing, people who inject drugs, harm reduction, South Africa, decentralized care, implementation research

## Abstract

People who inject drugs (PWID) and people who use drugs (PWUD) bear a disproportionate burden of hepatitis C virus (HCV) infection globally. In South Africa, HCV testing and treatment remain limited outside externally funded projects. This study investigated the implementation feasibility of assisted HCV self-testing (HCVST) among PWID and PWUD in Johannesburg.

Between 12^th^ May 2023 and 28^th^ March 2024, participants were recruited to an implementation study across mobile harm reduction sites and a central clinic. Participants performed self-tests using either oral-fluid or blood-based HCV antibody rapid tests. Reactive results were followed by on-site venous sampling for confirmatory RNA testing and referral for direct-acting antiviral (DAA) therapy at a centralized facility. We describe HCV case-detection, care cascade progression, and behavioral risk factors associated with HCV reactivity using logistic regression.

Of 1,566 participants tested, 998 (63.7%) were HCV reactive. The median age was 31 years (IQR 28–35); 82.2% were male and 77.1% identified as PWID. Ever injecting drugs (OR 35.6, 95% CI 23.6–56.0), frequent injecting (≥ daily: OR 36.7, 95% CI 25.1–55.3), and recent needle sharing (OR 7.3, 95% CI 5.8–9.3) were the strongest predictors of HCV reactivity. Histories of incarceration were also independently associated with HCV reactivity (OR 3.2, 95% CI 2.6–4.0). Despite high self-testing acceptability, progression through the care cascade was limited: among 854 RNA-confirmed infections, only 147 (17.2%) were prioritized for treatment, with three participants achieiving sustained virologic response. Thematic analysis identified fear of needles, poor venous access, and structural barriers, notably centralized treatment delivery, as key impediments to linkage.

This study showed a high burden of HCV among PWID and PWUD in Johannesburg and demonstrates that assisted HCVST is acceptable. Centralized treatment models severely constrained linkage to care. Simplified delivery of treatment is critical in transforming diagnosis into cure.

## Introduction

An estimated 58 million people are living with chronic HCV worldwide [1]. Despite the availability of curative therapies, only 36% of those infected had been diagnosed between 2015 and 2022 and 20% have received treatment [2], highlighting substantial disparities in access to testing and care. If untreated, HCV can lead to severe liver diseases, including cirrhosis and hepatocellular carcinoma, resulting in considerable morbidity and mortality [1]. People who Inject Drugs (PWID) and People who Use Drugs (PWUD) are disproportionately affected by HCV due to behaviours and circumstances that increase their risk of exposure [3, 4], for example sharing needles, and structural challenges such as criminalisation that hinder access to health services.

In many countries, including in South Africa, the HCV epidemic is concentrated amongst PWID, with small studies estimating prevalence of 84% in Pretoria against a general population background estimate of less than 1% [5, 6]. As in other settings, structural barriers to healthcare access increase vulnerability in South Africa, and outside of externally funded projects, HCV testing and treatment is not available. Although direct-acting antiviral (DAA) therapies can cure over 95% of individuals in many settings [17, 19] this potential is not realised domestically: testing and treatment remain limited. In South Africa, DAAs are not registered for general use and are accessible only via section 21 special authority applications through South African Health Products Regulatory Authority(SAHPRA) [15,18]. Hepatitis C testing is often conducted only through centralized laboratories and is not routine even among high risk key populations, including PWID, reflecting broader systemic gaps in timely diagnosis and care [19, 20].

Recognizing these challenges, the World Health Organization (WHO) has called for innovative strategies, and recommends HCV self-testing (HCVST), to scale up HCV testing and treatment to achieve the 2030 elimination targets [1]. Guidelines from WHO strongly recommend offering self-testing for hepatitis C virus as an additional approach to HCV testing services [7]. Prior evidence has shown that assisted self-testing at decentralised sites is an effective strategy for increasing HIV testing uptake among PWID [8,9]. Harm reduction sites, many of which already provide HIV testing, counselling, and linkage services, may offer promising platform for integrating and scaling up HCV testing modalities. Self-testing has demonstrated considerable benefits in expanding access to diagnostic services among marginalized and hard to reach populations, particularly in South Africa where HIV self-testing has been effectively scaled up [21]. If such sites can demonstrate effectiveness in identifying HCV positive individuals and facilitating timely linkage to confirmatory testing and treatment, they could serve as critical hubs for reaching high risk and underserved populations.

Here we assess the results of implementing HCVST among PWID and PWUD in mobile harm reduction sites across Johannesburg to explore the implementation feasibility of HCVST.

## Methods

### Ethics statement

Ethical approval for the study was obtained from the University of the Witwatersrand Human Research Ethics Committee (220306), South African Health Products Regulatory Authority (MD20220801), South African National Clinical Trial Registry unique identification number for the registry is DOH-27-122022-4828, and World Health Organization Ethics Review Committee (0003847). All participants provided written informed consent prior to participation.

### Study design and setting

We recruited participants to an implementation study between 12^th^ May 2023 and 28^th^ March 2024 to evaluate the feasibility of HCVST among PWID, PWUD, men who have sex with men (MSM), and sex workers in Johannesburg, South Africa. The study was implemented by Ezintsha (University of the Witwatersrand) in collaboration with harm reduction programs operating under the JabSmart initiative. Activities were based at a fixed harm reduction site in Yeoville and through mobile outreach services that operated across Johannesburg’s seven administrative regions, targeting high-risk community hotspots.

### Participants

Adults aged ≥18 years accessing harm reduction or HIV testing services at participating sites were eligible for study inclusion. Recruitment occurred through convenience sampling during routine clinic visits and mobile outreach sessions. Key population identity was determined through self-report and verified based on existing engagement with harm reduction services.

### Intervention overview

Participants were provided with an HCV self-test to perform on site under silent observation by trained study staff. Two test types were used: the First Response HCV card test (Premier Medical Corps Ltd., India) and the OraQuick HCV rapid antibody test (OraSure Technologies Inc., USA). Both were repackaged by manufacturers as self-tests with translated, manufacturer-approved instructions for use. Participants interpreted their own results, which were then verified by the observer.

Participants with reactive HCV self-tests were offered confirmatory HCV RNA polymerase chain reaction (PCR) testing via venous blood sampling. Samples were transported to a certified Johannesburg laboratory for RNA confirmation and ancillary testing, including aspartate aminotransferase (AST) and platelet counts for liver fibrosis assessment. Turnaround time for results was approximately two days. Participants were provided printed results and offered treatment initiation through the Yeoville clinic once direct-acting antivirals (DAAs) became available midway through the study (December 2023). Participants testing positive prior to this date were re-contacted once treatment was available.

DAA initiation occurred at the Yeoville clinic under the supervision of a study physician. Prrioritsed participants (HCV RNA positive, HIV negative, and hepatitis B negative) initiated DAA therapy through directly observed treatment (DOT). Those who declined clinic referral were revisited by field teams to receive their results and counselling. Treatment completion and sustained virologic response (SVR) were documented through medical chart abstraction.

### Variables

Data were collected using REDCap electronic forms and supplemented by clinical records. The primary outcome was implementation feasibility defined in two ways; the first was HCV self-test case detection, defined as the proportion of participants testing HCV reactive. The second was progression along the HCV cascade: (1) self-test reactivity, (2) confirmatory RNA testing, (3) treatment initiation, and (4) treatment completion in those eligible for each step.

Sociodemographic variables included age, gender, education, and key population category. Behavioral risk factors were captured using a structured questionnaire assessing injecting practices, needle sharing, incarceration, sexual behaviors, tattooing, medical procedures, and household blood exposure.

Acceptability of self-testing was assessed through five Likert-scale items (ease, convenience, privacy, trust, and comfort), which were combined into a composite score (Cronbach’s α = 0.67). Scores were categorized as low (<3.0), moderate (3.0–<4.0), or high (4.0–5.0).

### Statistical analysis

Data was analysed using R. Participant study throughput was described. The primary study outcome implementation feasibility in terms of case-detection was described through the proportion testing HCV reactive using the HCV self-test with binomial exact confidence interval. Sociodemographic variables of all testing participants were described using frequency statistics, as well as characteristics of the tests conducted (site type, test used, and overall acceptability), and compared by HCV self-test reactivity. Given high prevalence of HCV in this population, additional risk factors hypothetically associated with HCV prevalence in this group were explored and their association with HCV self-test reactivity investigated through univariate logistic regression. Missingness was described throughout.

To firther explore implementation feasibility, the HCV care cascade was graphed to describe progression from assisted self-testing through to confirmatory testing and treatment initiation (where eligible), with proportions and drop-offs reported at each stage. Cascade steps were: (1) HCV self-test reactive, (2) PCR uptake, (3) PCR confirmation, if eligible (3) treatment initiation and (4) treatment completion.

Given very low treatment initiation and completion proportions of those eligible for treatment we thematically analysed reasons given for non-completion of confirmatory tests. We then conducted univariate analysis by testing site (outreach versus centralised facility). Given DAAs were only available for initiations at the centralised Yeoville facility, we hypothesized that those who had attended for a test at the centralised service may be more likely to initiate and complete their DAA course.

## Results

### Participant throughput

1569 participants enrolled to test for HCV of which 1566 had a recorded reactive or negative result and were included in analysis (2 were missing, and 1 was “Other”); 233 tested at the central Yeoville clinic and 1333 at outreach sites using the van. 998 people tested reactive using the HCVST kit (63.7% case-detection, 95% CI 61.2% to 66.0%) (Fig 1).

**Fig 1.**
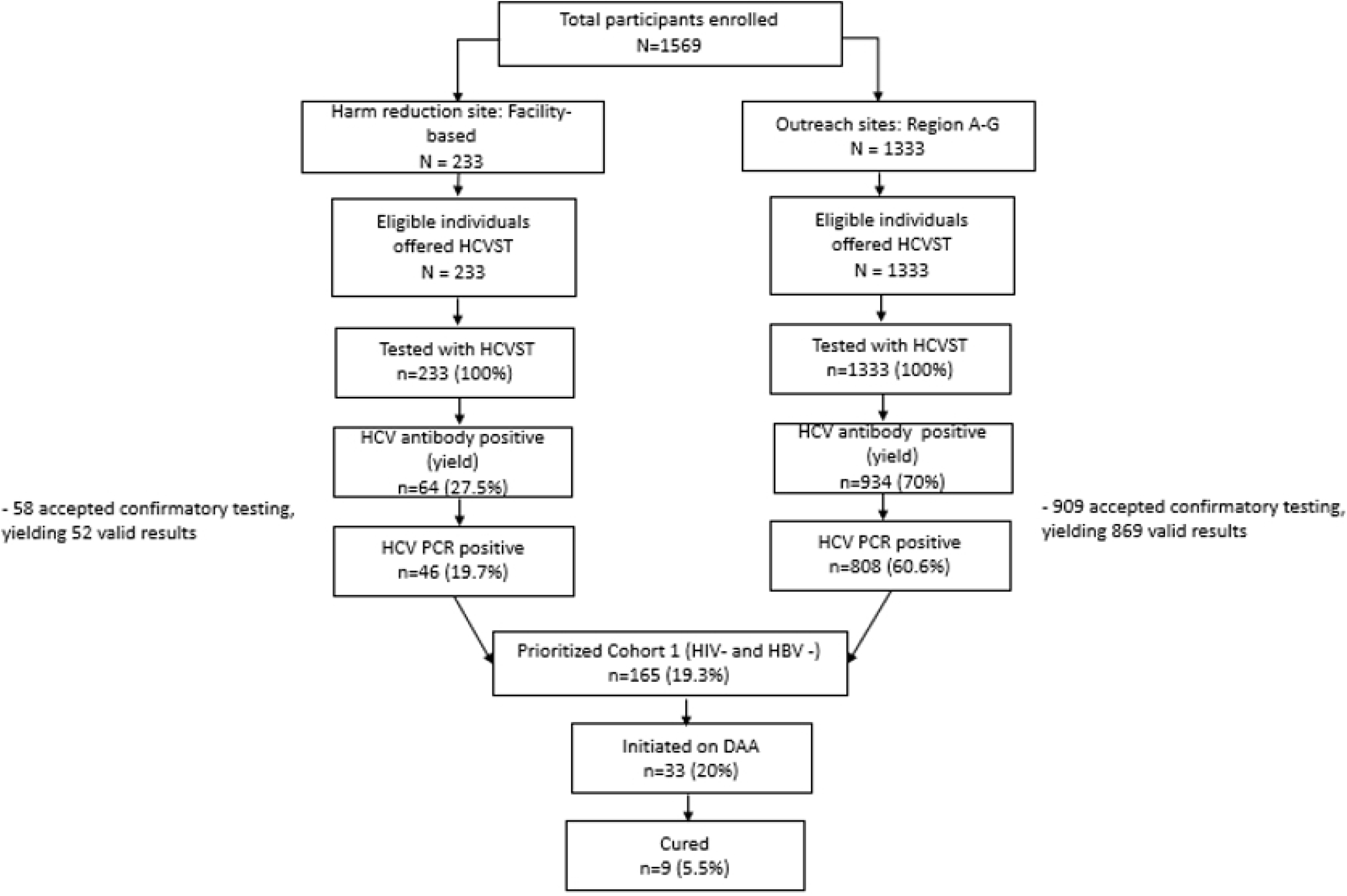
Study participant flow

### Study Population

The median participant age was 31 years (IQR 28–35 years); almost two-thirds of the participants were aged 25–34 years (61.8%, 969/1566) and the majority were male (82.7%,1286/1556). Most participants (77.1%, 1208/1566) were PWID while only 3.6% (56/1566) were MSM. Most participants (86.4%, 1354/1566) had a secondary level of education (grade 8–12) (**Table 1**).

**Table 1.** Participant characteristics.

| Socio-demographic factors and testing characteristics* | Total<br>N = 1566<br>(%) | HCVST<br>negative<br>n = 568<br>(36.3%) | HCVS<br>T<br>reactiv<br>e<br>n = 998<br>(63.7%<br>) | p-<br>value<sup>†</sup> |
| --- | --- | --- | --- | --- |
| **Age** |  |  |  |  |
| Median age in years (IQR) | 31 (28 to 35) | 33 (28 to 38) | 31 (28 to 34) | <0.001<sup>1</sup> |
| 18-24 | 123 (7.9%) | 61 (10.7%) | 62 (6.2%) | <0.001 |
| 25-34 | 967 (61.7%) | 275 (48.4%) | 692 (69.3%) |  |
| 35-44 | 418 (26.7%) | 189 (33.3%) | 229 (22.9%) |  |
| 45+ | 58 (3.7%) | 43 (7.6%) | 15 (1.5%) |  |

| Gender identity<sup>2</sup> |  |  |  |  |
| --- | --- | --- | --- | --- |
| Male | 1288<br>(82.2%) | 367<br>(64.6%) | 921<br>(92.3%) | <0.001 |
| Female | 273 (17.4%) | 196<br>(34.5%) | 77<br>(7.7%) |  |
| Educational level |  |  |  |  |
| ≤Grade 7 primary schooling level | 44 (2.8%) | 23<br>(4.0%) | 21<br>(2.1%) | <0.001 |
| ≥Grade 8 primary schooling to ≤High School | 1353<br>(86.4%) | 452<br>(79.6%) | 901<br>(90.3%) |  |
| ≥Post High School Institution, University and University plus | 169 (10.8%) | 93<br>(16.4%) | 76<br>(7.6%) |  |
| Key population identity<sup>3</sup> |  |  |  |  |
| PWID | 1208<br>(77.1%) | 231<br>(19.1%) | 977<br>(80.9%) | <0.001 |
| PWUD | 159 (10.2%) | 138<br>(86.8%) | 21<br>(13.2%) | <0.001 |
| MSM | 56 (3.6%) | 56<br>(100.0%) | 0<br>(0.0%) | <0.001 |
| SW | 143 (9.1%) | 143<br>(100.0%) | 0<br>(0.0%) | <0.001 |
| HIV status |  |  |  |  |
| Negative | 636 (40.6%) | 159<br>(28.0%) | 477<br>(47.8%) | <0.001 |
| Positive | 524 (33.5%) | 102<br>(18.0%) | 442<br>(44.3%) |  |
| Unknown | 406 (25.9%) | 327<br>(57.6%) | 79<br>(7.9%) |  |
| Site type |  |  |  |  |
| Outreach setting (van) | 1333<br>(85.1%) | 399<br>(70.2%) | 934<br>(93.6%) | <0.001 |
| Yeoville Clinic | 233 (14.9%) | 169<br>(29.8%) | 64<br>(6.4%) |  |
| Test type |  |  |  |  |
| Oral-saliva test | 688 (43.9%) | 197<br>(34.7%) | 491<br>(49.2%) | <0.001 |
| Blood-based test | 878 (56.1%) | 371<br>(65.3%) | 507<br>(50.8%) |  |
| Intervention acceptability |  |  |  |  |
| High intervention acceptability (mean score ≥ 4.0) | 1470<br>(93.0%) | 546<br>(96.1%) | 924<br>(92.6%) | 0.057<sup>4</sup> |
| Moderate intervention acceptability (mean score ≥ 3.0, < 4.0) | 65 (4.2%) | 17<br>(3.0%) | 48<br>(4.8%) |  |
| Low intervention acceptability (mean score < 3.0) | 3 (0.2%) | 0 (0.0%) | 3<br>(0.3%) |  |

\*Missingness described: Gender (n=5 reported as other, 0.3%), Intervention acceptability (n=28 missing responses across all five items, 1.8%).
† Chi-square test unless otherwise reported
<sup>1</sup> Wilcoxon rank-sum test
<sup>2</sup> Of which 4 male and 3 female were assigned a different gender at birth (4 trans-male and 3 trans-female.)
<sup>3</sup> Row percentages rather than column percentages shown here, as each KP identity is treated as a distinct group: hence separate chi square tests.
<sup>4</sup> Low and moderate combined for the purposes of the chi square test.

Most participants were tested at outreach sites (85.1%, 1333/1566) compared to the the central Yeoville clinic (14.9%, 233/1566). Intervention acceptability was high (93.0% high acceptability score; 1470/1566).

Participants who tested reactive tended to be slightly older (particularly in the 25–34 age group; 69.3% reactive compared to 48.4%, p<0.001), predominantly male (92.3% comapared to 64.6% p<0.001, and predominantly self-identified as PWIDs (80.9% compared to 19.1%, p<0.001). They were also more likely to be tested in outreach settings (93.6% compared to 70.2%, p<0.001). Differences in the overall acceptability of the intervention between those testing reactive and those testing negative were marginal (high 92.6% compared to 96.1%, p=0.057).

There was a high burden of HIV in the entire population (33.5%, 524/1566) attesting to the vulnerability of this client population. In people with HCV, there was also a large amount of HIV coinfection (47.8%, 477/998). This appeared greater than people testing HCV negative (28.0%, 159/568), but the large amount of missing data especially amongst those testing HCV negative make direct comparisons challenging.

### Prevalence of HCV risk factors

The strongest predictor of HCV reactivity was history of injecting drug use. Participants who reported ever injecting had 35-fold higher odds of being HCV reactive compared to those who have never injected (OR 35.6, 95% CI: 23.6-56.0, *p*<0.001) (Table 2).

**Table 2.**
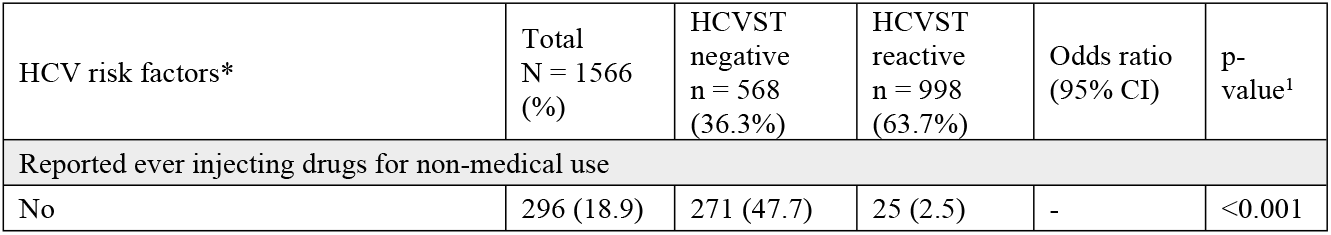
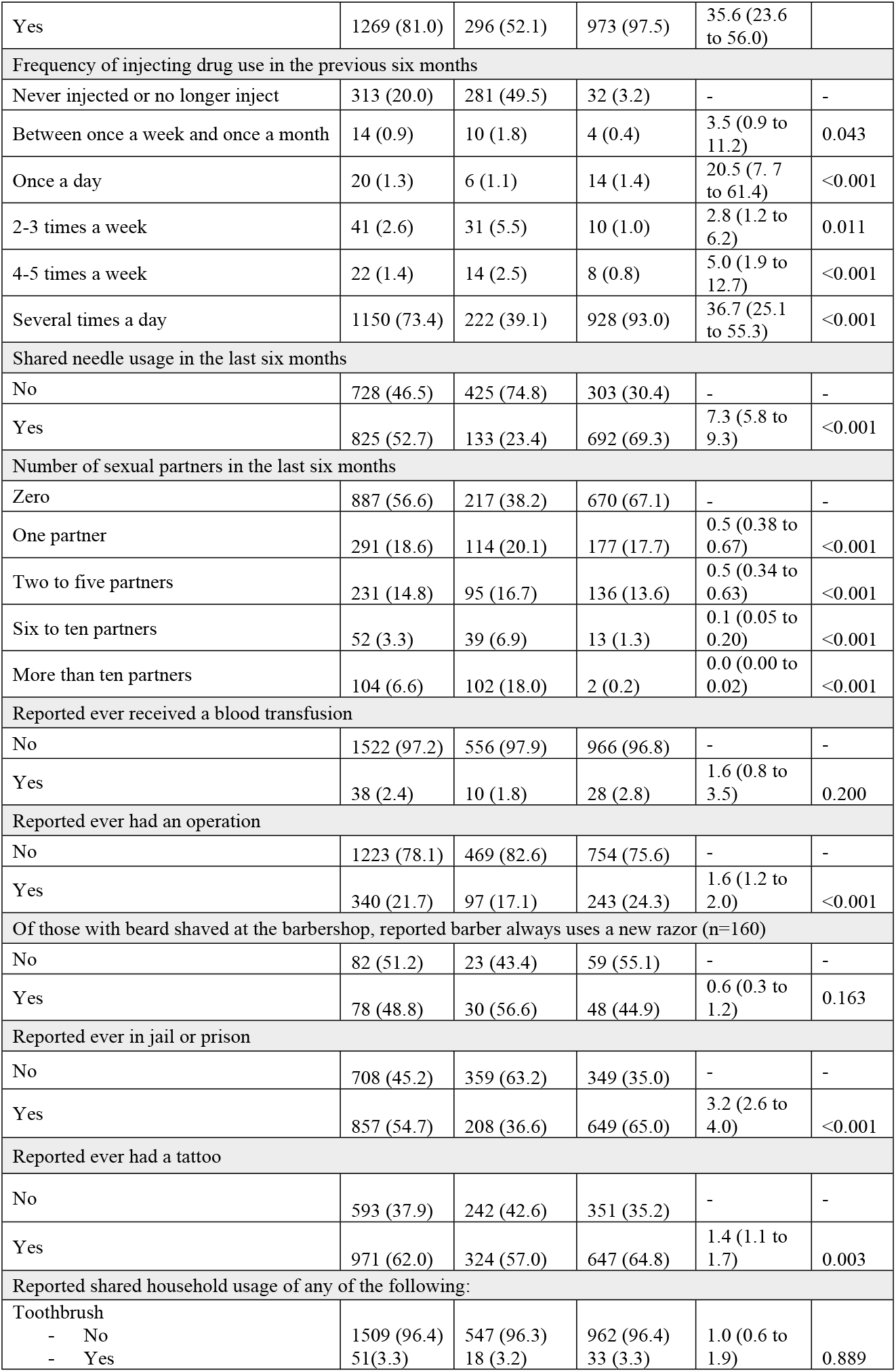
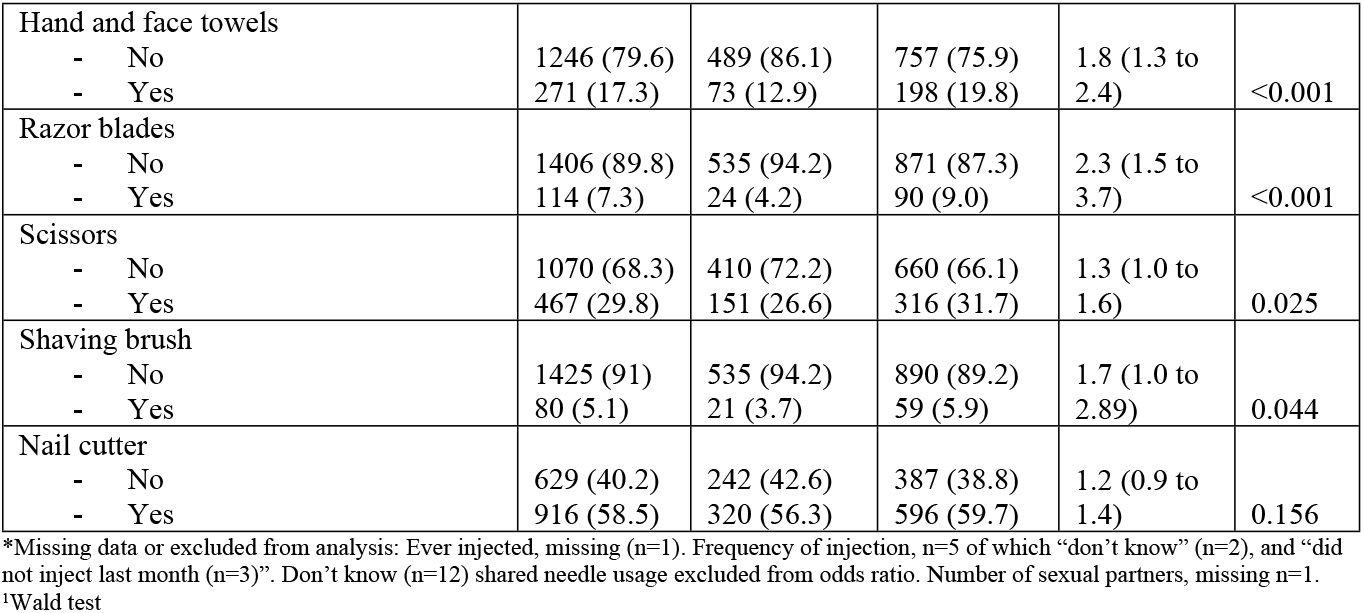
HCV risk factor prevalence.

Frequency of injecting was also a critical risk factor: those injecting several times a day had higher prevalence of HCV reactivity (93.0%) with 36 times the odds of HCV (OR 36.7, 95% CI: 25.1-55.3, p<0.001) compared to those who do not inject. The risk remains elevated among those injecting 4-5 times weekly with five times the odds of HCV (OR 5.0, 95% CI: 1.9-12.7, p<0.001) and 2-3 times weekly almost three times the odds (OR 2.8, 95% CI: 1.2-6.2, p=0.011). Needle sharing was another significant risk factor. Participants who reported sharing needles in the past six months had over 7 times the odds of testing reactive (OR 7.3, 95% CI: 5.8-9.3, p<0.001). By contrast, sexual risk factors showed a negative association with HCV reactivity; the more sexual partners, the reduced odds of HCV reactivity indicating that sexual behaviour appears less relevant than injecting practices for HCV transmission. Histories of incarceration were highly prevalent with over three times the odds of HCV infection: over half of participants (54.7%) reported ever being in jail or prison and this group had 3 times higher odds of testing reactive (OR 3.2, 95% CI: 2.6-4.0, p<0.001).

### HCV cascades

The HCV treatment and care cascade highlights a major bottleneck between lab detection and treatment initiation, where most confirmed cases did not transition into care. Of the 1566 participants who used the self-test, 63.7% were antibody positive, and almost all these people (97.1%, 969/ 998) completed confirmatory testing. Among these, most (88.1%, 854/969) were PCR positive (viraemic). However, under a fifth (17.2%, 147/854) of those with confirmed viraemic infection were prioritized for treatment. Of those prioritised for treatment, only a quarter (25.9%, 38/147) were able to initiate therapy, and under half (42.1%, 16/38) completed treatment. Ultimately only 3 participants achieved sustained virologic response (Fig 2).

**Fig 2.**
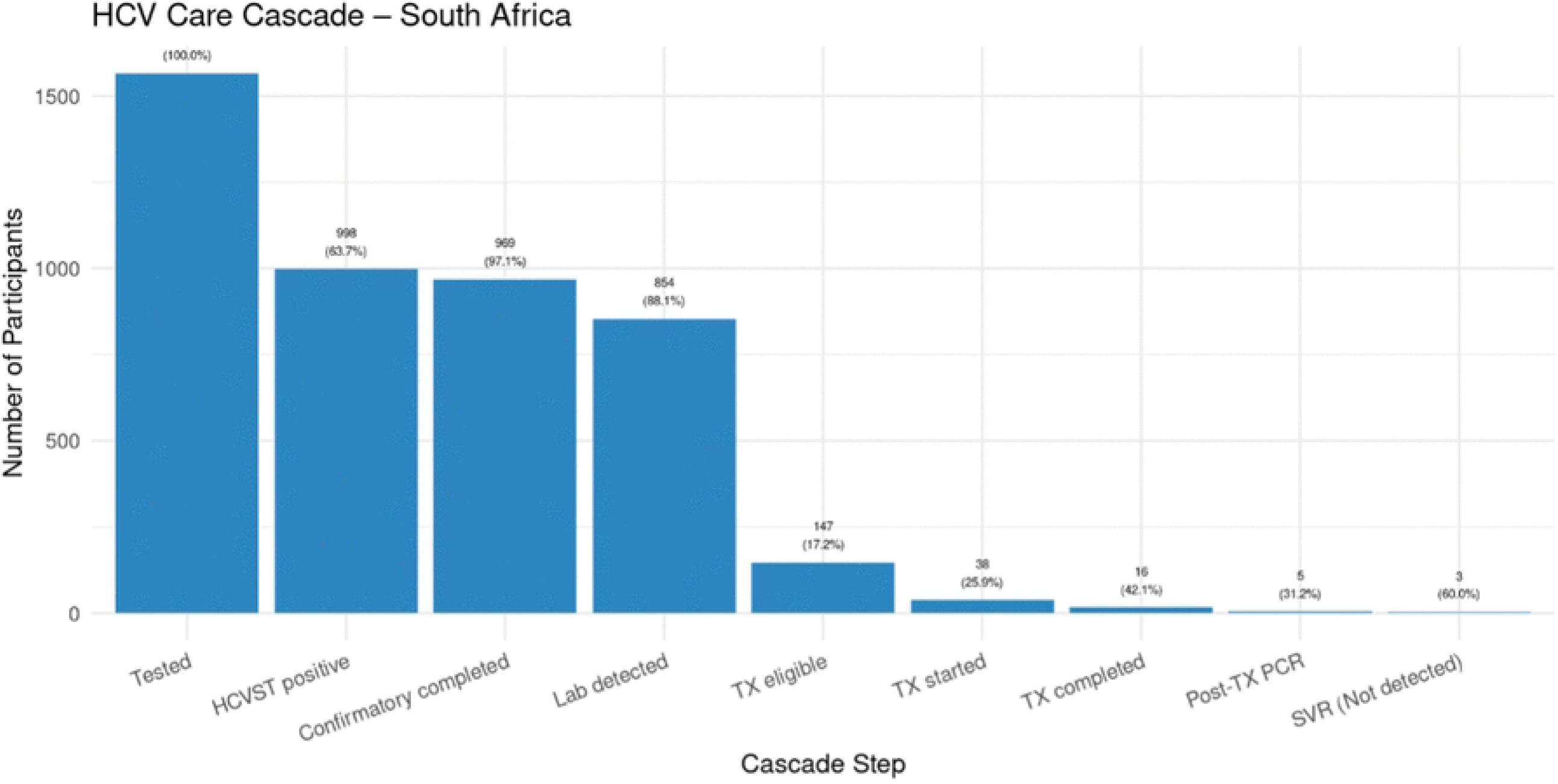
HCV Care Cascade

### Exploratory analysis of the HCV care cascade

Given very low treatment initiation and completion proportions of those eligible for treatment we thematically analysed reasons given for non-completion of confirmatory tests, hypothesizing these identified barriers may present across cascade steps. Among 28 participants who provided reasons for not completing confirmatory testing, their responses could be grouped into four predominant themes. The most common being afraid of needles or refusal to provide a blood sample (11/28; 39.3%), with participants citing fear of needles, discomfort, or unwillingness to have blood drawn. The second most frequent reason was difficulty in obtaining a blood sample due to poor or inaccessible veins (10/28; 35.7%). However, there were also implementation challenges identified. Prior to on-site blood draw for PCR, PCR testing was administered only at the central Yeoville clinic. One-tenth of participants (10.7%, 3/28) could not wait to be transported to this centralised facility or were in a hurry. Finally, withdrawal of the study accounted for remaining reasons (2/28; 7.1%), where participants promised to return but did not.

Given DAAs were only available for initiations at the centralised Yeoville facility, we had hypothesized that those who had attended for a test at the centralised service may be more likely to initiate and complete their DAA course. However, only 5 people who tested reactive and were eligible for treatment had tested at the central clinic site and none of those started treatment, therefore we were unable to run regression analyses on these.

## Discussion

This study demonstrated an alarmingly high burden of hepatitis C virus among people who inject drugs (PWID) and people who use drugs (PWUD) in Johannesburg, South Africa. Nearly two-thirds (63.7%) of participants had reactive HCV self-test results, rising to 97.5% among those who had ever injected drugs, highlighting that HCV infection is likely widespread in this population. Frequent injection, receptive needle sharing and incarceration emerged as strong predictors of HCV prevalence, while sexual behaviours and non-injection exposures contributed little. HIV/ HCV coinfection was also very high in this population, pointing to a highly vulnerable population in a situation of acute epidemic. These findings reinforce global evidence that unsafe injecting practices remain the primary driver of HCV transmission among PWID and that the explicit vulnerabilities affecting PWID and PWUD, including criminalization and incarceration, perpetuate transmission cycles [4].

Despite the high uptake and acceptability of HCV self-testing, progression through the care collapsed at the point of treatment initiation. Of 854 participants with RNA confirmed infection, only 17% were prioritised for treatment and just three achieved sustained virologic response. This highlights a fundamental structural challenge: while HCV testing was decentralized and community based, treatment was available only in a centralized clinic using directly observed treatment protocols. This disconnect between diagnosis and care delivery undermines the potential of outreach based testing models to translate diagnosis into cure.

Our findings are consistent with studies from other resource limited settings, which demonstrate that facility based models alone are not suited for reaching underserved and criminalized populations such as PWID and PWUD. Simplified point-of-care HCV models in Cape Town for example, achieved improved linkage when confirmatory testing and treatment were co-located within harm reduction sites rather than requiring referral to clinics [6].

Decentralization has been shown to be beneficial in international settings [11]. Australian model of hepatitis C decentralized care has shown excellent outcomes of HCV treatment [12]. Lessons from HIV programs, particularly the use of decentralized service delivery, community based and differentiated care models are highly relevant for addressing gaps in the HCV care cascade [9,14].

That linkage faltered because confirmatory testing required venipuncture and clinic attendance is indicative of broader health systems and access issues beyond HCV that must be addressed. As indicated in a recent systematic review on linkage following HIVST, linkage was high and aligned with the standard linkage rates [16]. Further, evidence has shown that home-based assessment or initiation of care can enhance linkage after self-testing [22]. In Lesotho, peer-led and community-based HIV service delivery models (eg. peer educators) have been used in other HIV care contexts, though linkage data post self-testing remain limited [23]. Given the challenges PWID/PWUD face in South Africa, efforts are needed to improve rates of healthcare utilization, including HCV testing and care. While self-testing can overcome many challenges to access, it can only achieve impact with more diverse adaptations to the delivery of HCV prevention and treatment services.

This study has several limitations. Treatment was initially prioritized for participants who were HIV negative and hepatitis B negative, leaving a substantial proportion of coinfected individuals untreated when DAAs became available. The lack of recency assays limited our ability to characterize ongoing transmission versus longstanding infection. The observational design and missing data at various points in the cascade may have introduced bias in estimates of linkage and treatment initiation.

## Conclusion

This study demonstrates both the promise and the limitations of assisted HCV self-testing within current systems. To meaningfully reduce transmission and morbidity, HCV programs in South Africa must move beyond centralized models and align with the lived realities of PWID, ensuring that testing becomes not just an entry point, but a pathway to cure.

## Data Availability

All relevant data are within the paper and its Supporting Information files.

